# Survival of vaccine-induced human milk SARS-CoV-2 IgG and IgA immunoglobulins across simulated human infant gastrointestinal digestion

**DOI:** 10.1101/2021.06.17.21259021

**Authors:** Myrtani Pieri, Vicky Nicolaidou, Irene Paphiti, Spyros Pipis, Kyriacos Felekkis, Christos Papaneophytou

## Abstract

Four vaccines have been approved to date by the European Medicines Agency for the management of the COVID-19 pandemic in Europe, with all four being targeted to adults over 18 years of age. One way to protect the younger population such as infants or younger children until pediatric vaccines are licensed is through passive immunity via breastfeeding. Recent evidence points to the fact that human milk contains immunoglobulins (Ig) against the SARS-CoV-2 virus, both after natural infection or vaccination, but it is not known whether these antibodies can resist enzymatic degradation during digestion in the infant gastrointestinal (GI) tract or indeed protect the consumers. Here, we describe our preliminary experiments where we validated commercially available ELISA kits to detect IgA and IgG antibodies in human milk from two lactating mothers vaccinated with either the Pfizer/BioNTech or the Astra Zeneca vaccine, and the effect of a static *in vitro* digestion protocol on the IgA and IgG concentrations. Our data, even preliminary, provide an indication that the IgA antibodies produced after vaccination with the Pfizer/BioNTech vaccine resist the gastric phase but are degraded during the intestinal phase of infant digestion, whereas the IgGs are more prone to degradation in both phases of digestion. We are in the process of recruiting more individuals to further evaluate the vaccine induced immunoglobulin profile of breastmilk, and the extent to which these antibodies can resist digestion in the infant GI tract.

## 1. Introduction

The ongoing novel coronavirus disease 2019 (COVID-19) outbreak, caused by the severe acute respiratory syndrome coronavirus 2 (SARS-CoV-2), has posed global public health concern due to its high dispersion rate influencing public health, society, and the economy across the world (Sharma et al., 2020; Zhu et al., 2020). To date, four vaccines have been approved to target COVID-19 by the European Medicines Agency (EMA) for use in Europe, two employing mRNA technology (BNT162b2 mRNA by Pfizer-BioNTech and mRNA-1273 by Moderna) and two employing recombinant adenovirus technology (ChAdOx1 nCoV-19 by Oxford/AstraZeneca and Ad26.COV2.S by Johnson & Johnson) (Yan et al., 2021). However, none of these vaccines are authorized or currently under investigation for use in infants or very young children. Even though young children are usually only mildly affected by COVID-19, there have been reports of the appearance of a multisystem inflammatory syndrome in this population, linked to SARS-CoV-2 infection (Dufort et al., 2020).

For breast-fed infants, human milk is a source of various nutrients (e.g., proteins, peptides) and bioactive components that promote neonatal growth and protect from viral and bacterial infection (Kompaneets et al., 2020). In terms of antibodies, human milk contains an array of immunoglobulins (Igs), including IgA, secretory IgA (SIgA), IgM, secretory IgM (SIgM), and IgG [see (Czosnykowska-Łukacka et al., 2020a) and references cited therein].

IgA is the most abundant antibody isotype (∼90% of the total Ig) in human milk, (Hurley & Theil, 2011; Palmeira & Carneiro-Sampaio, 2016), mostly found as SIgA; a dimer bound to a secretory component (Goldman et al., 2011). IgA antibodies in human milk protect the infant by eliminating invading pathogens on mucosal surfaces or systemically (Palmeira & Carneiro-Sampaio, 2016), while specific IgA antibodies have been detected in human milk of mothers previously infected by SARS-CoV, human immunodeficiency virus (HIV) and respiratory syncytial virus (Palmeira & Carneiro-Sampaio, 2016; Robertson et al., 2004).

It has been previously demonstrated that human milk from mothers who have been infected with SARS-CoV-2 contains antibodies against the virus, and that these antibodies also demonstrate a neutralizing ability (Fox et al., 2020b; Pace et al., 2021; van Keulen et al., 2020). The same has been shown for mothers vaccinated with the mRNA vaccine technology, where SARS-CoV-2-specific IgG and IgA antibodies seem to appear in breastmilk soon after the first dose and reach maximum levels about 2 weeks after the second dose (Kelly et al., 2021; Pace et al., 2021; Perl et al., 2021). A number of pre-prints have also shown that even though both IgA and IgG antibodies appear in milk after vaccination, IgG seems to be the prominent isotype reported in the majority of studies but results are still inconclusive (Baird et al., 2021; Bertrand et al., 2021; Esteve-Palau et al., 2021; Fox et al., 2021; Friedman et al., 2021; Golan et al., 2021).

Whichever the antibody profile secreted in milk upon vaccination, it is understood that in order for any such antibodies to play a role in immunoprotection, Igs must resist proteolytic degradation throughout digestion and remain intact and functional in order to bind pathogens in the digestive system and block that route of infection. Also, to provide a more systemic protection, these antibodies, must be absorbed into the bloodstream of infants (Demers-Mathieu et al., 2019). Importantly, one study demonstrated that the vaccine-elicited Ig profile in milk after COVID-19 mRNA-based vaccination (Pfizer/BioNTech and Moderna) is IgG-dominant and lacks secretory antibodies which is the Ab class that is highly stable and resistant to enzymatic degradation in all mucosae (Fox et al., 2021). To date, no study has examined if the vaccine-induced antibodies withstand the infant digestion process.

In this preliminary case study involving two lactating mothers, we first validated the accuracy of a commercially available ELISA assay for the determination of anti-SARS-CoV-2 IgAs and IgGs in breastmilk. We then employed this kit to evaluate the Ab response in milk after vaccination with both doses of either the Pfizer/BioNTech or the Astra Zeneca vaccine. We are also the first to evaluate the effect of digestion on the antibodies elicited by the Pfizer/BioNTech vaccine by employing a static *in vitro* protocol of infant digestion.

## 2. Materials and methods

### 2.1 Study participants

Two lactating women were recruited in the present study that had no diagnosis of laboratory-confirmed COVID-19 in the past and were scheduled to receive a COVID-19 vaccine. Participants gave informed consent, and all procedures were approved by the Cyprus National Bioethics Committee (EEBK/EΠ /2021/24). Participant A received the two-dose adenovirus-vector Astra Zeneca vaccine (ChAdOx1 nCoV-19), 8 weeks apart. Participant B received the two-dose Pfizer/Biotech vaccine (BNT162b2 mRNA) 3 weeks apart as per standard protocols in Cyprus at the time. Additional characteristics of study participants and their infants are presented in **Table 1**.

**Table 1.** Maternal and infant characteristics.

|  | **Mother A** | **Mother B** |
| --- | --- | --- |
| Maternal Age | In their 30's | In their 30's |
| Post-partum | 11 months | 7 months |
| Type of vaccine | Astra Zeneca | Pfizer/BioNTech |

Control milk samples were obtained before vaccination and used as controls to establish positive cutoff values for each assay. Also, samples were collected 1 day before the second dose and 3 weeks after the second dose to determine antibody levels. All samples were divided into 1.5-mL aliquots and stored at -80 °C.

### 2.2 SARS-CoV-2 IgA and IgG ELISA validation

ELISA kits specific for the determination of IgA and IgG immunoglobulins against SARS-Cov-2 were obtained from Abcam (Cat numbers: ab277285 for IgG and ab277286 for IgA). The spectrophotometric ELISA assays were monitored with a microplate reader (PerkinElmer, Waltham, MA). The concentration of both IgA and IgG positive control samples (provided by the manufacturer) was determined using the Bradford method (Bradford, 1976), while bovine serum albumin (BSA) was used to create a standard curve. Two human milk samples (500 μL each) were supplemented with 200 μg/mL IgA or IgG against SARS-CoV-2. The specific SARS-CoV-2 IgA and IgG ELISA assays were validated for accuracy and precision as previously described (Andreasson et al., 2015) with some modifications. All ELISA tests were performed for the two human milk samples on the same day with two dilutions and in triplicate. The accuracy of the assays was evaluated using the % error from the following equation % Error = (V_A_ – Vo)/V_A_ × 100, where V_A_ is the known value and Vo represents the observed value. The precision of the assays was evaluated using the % coefficient of variation (CV), as follows: % CV = Standard deviation (SD)/ Average × 100. The recovery of the assays was evaluated via the equation % Recovery= (M_S_-M_N_)/T_S_ ×100 (Andreasson et al., 2015), where M_S_ is the measured concentration of the spike sample, M_N_ is the measured concentration of the neat samples, and T_S_ is the theoretical concentration of the spiked sample. Unspiked milk was used as a blank in these experiments.

### 2.2 *In vitro* digestion of human milk

A two-step *in vitro* infant GI digestion method that consists of a 60-min of gastric phase and a 120-min of intestinal phase as previously described by (Lueangsakulthai et al., 2020) and (Nguyen et al., 2015) was used in this study with slight modifications. Briefly, for gastric digestion, milk samples (1 mL) were mixed with 500 μL of simulated gastric fluid consisting of 0.15 M NaCl, pH 4.0 supplemented with 0.3 mg pepsin (22.75 U/mg total protein). The pH of each sample was re-adjusted to 4.0 ± 0.05 with 1 N HCl. The samples were then incubated at 37 °C with continuous shaking at 300 rpm for 60 min. When the incubation time for the gastric phase of digestion elapsed, a 300-μL aliquot was removed and stored at −80 °C. The remaining samples continued to the simulated intestinal digestion. The pH of each sample was adjusted to 8.0 by adding 1 N NaOH. Bile salts (0.4 mg) (Sigma-Aldrich, St. Louis, MO, USA) were added to make a final concentration of 2 mM bile salts. Porcine pancreatin (0.1 mg; 3.45 U of pancreatin/mg of total protein) (8 × USP, Sigma-Aldrich) was added to the mixture and the pH was readjusted to pH 8.0 ± 0.05. The mixture was incubated at 37 °C with shaking at 300 rpm in for 120 min. At the end of the incubation, samples were stored at −80 °C until further analysis.

### 2.3 Determination of IgA and IgG levels

Milk samples were quickly thawed in a water bath at 37°C and centrifuged at 1300 x g for 20 min at 4°C, fat was removed and supernatant transferred to new tubes and centrifuged again under the same conditions to remove residual fat and cells. Fat-free milk was aliquoted and stored at -80°C until testing. For the evaluation of the levels of IgA and IgG specific to SARS-CoV-2 spike protein in human milk, commercially available ELISA kits from Abcam that were developed for use in human plasma and serum (ab277285 for IgG and ab277286 for IgA) were used with modifications as mentioned above. Milk was either undiluted or diluted 1:2 using assay buffer (provided by the manufacturer) (for IgA) or 1:2 (or 1:4) using assay buffer supplemented with 1% w/v BSA (for IgG) and added to the plate. Antibody levels were reported as Units (U) using the following equation (that was provided by the manufacturer) Units= ((Sample Absorbance x 10 / Cut-off) × DF)), using as cut-off the mean absorbance values of milk samples before vaccination, where DF is the dilution factor. Samples containing >11 Units of IgA or IgG were considered as positive.

### 2.4 Statistical analysis

Unless otherwise stated, data are reported as means ± standard deviation (SD). The concentrations of antibodies before and after vaccination were compared with paired Student’s t-tests. One-way ANOVA with Tukey’s multiple comparison test was used to compare percentage stability of each antibody before and after digestion. All reported p-values were two-tailed and p-values < 0.05 were considered statistically significant. Statistical analysis was performed using GraphPad Prism (v.8.0, GraphPad Software Inc., San Diego, CA, USA).

## 3. Results

### 3.1 SARS-CoV-2 IgG and IgA ELISA method validation

In this work we used commercially available ELISA kits from Abcam (developed for use with serum and plasma samples) to evaluate the levels of SARS-Cov-2-specific IgA and IgG in human milk samples. Initially, the ELISA assays were validated using IgA and IgG spiked human milk samples (**Table 2**). In detail, two human milk samples spiked with either IgA or IgG specific to SARS-CoV-2 were analyzed within a single day with two dilutions for each sample and three replicates. The SARS-CoV-2 IgA and IgG ELISA assays were highly accurate with 5.17 % and 18.25 % error, respectively. The % CV (co-efficient variation) of SARS-Cov-2 IgA and IgG ELISA assays were 8.51 and 16.73%, respectively which meets the minimal validation requirements for assay precision (i.e., % CV < 30) according to (Andreasson et al., 2015). It should be noted that the addition of BSA to a final concentration of 10 mg/mL significantly increased (*p* <0.001) the recovery of spiked-IgG in human milk samples from 53.6 % to 83.75 % (data not shown) and decrease the assay error from ∼50 % to 18.25 %. Therefore, BSA was included in the subsequent ELISA experiments for the determination of IgG levels in human milk samples. On the contrary, the presence of BSA did not affect spike IgA recovery and assay % error (data not shown).

**Table 2.**
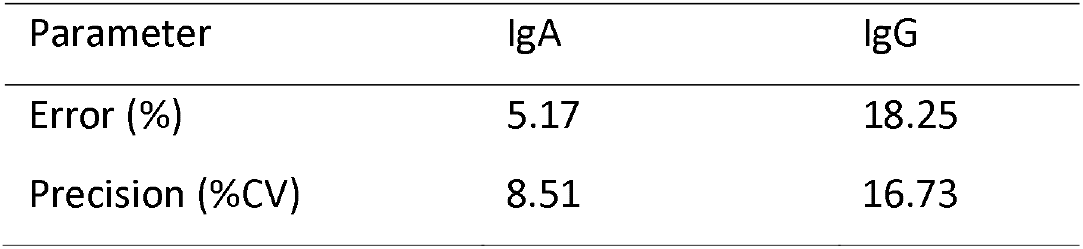
SARS-Cov-2 IgA and IgG ELISA method validation.

### 3.2 SARS-Cov-2 IgA and IgG levels in milk samples after vaccination with Pfizer/BioNTech and Astra-Zeneca Vaccines

We next compared the immune response to each vaccine type. Milk samples were obtained from one individual who received the Astra Zeneca vaccine and one who received the Pfizer/BioNTech vaccine. Samples were collected one day before the first dose, one day before the second dose and 3 weeks after the second dose. Fat-free milk samples were assayed in separate commercially available ELISAs measuring either IgA or IgG against SARS-CoV-2 (**Figure 1**). Milk samples were defined as IgA or IgG positive if the measured immunoglobulin units were greater than 11, the pre-vaccination (pre-vac) milk samples were used as a control (see methods, section *2.3* *Determination of IgA and IgG levels*).

**Figure 1.**
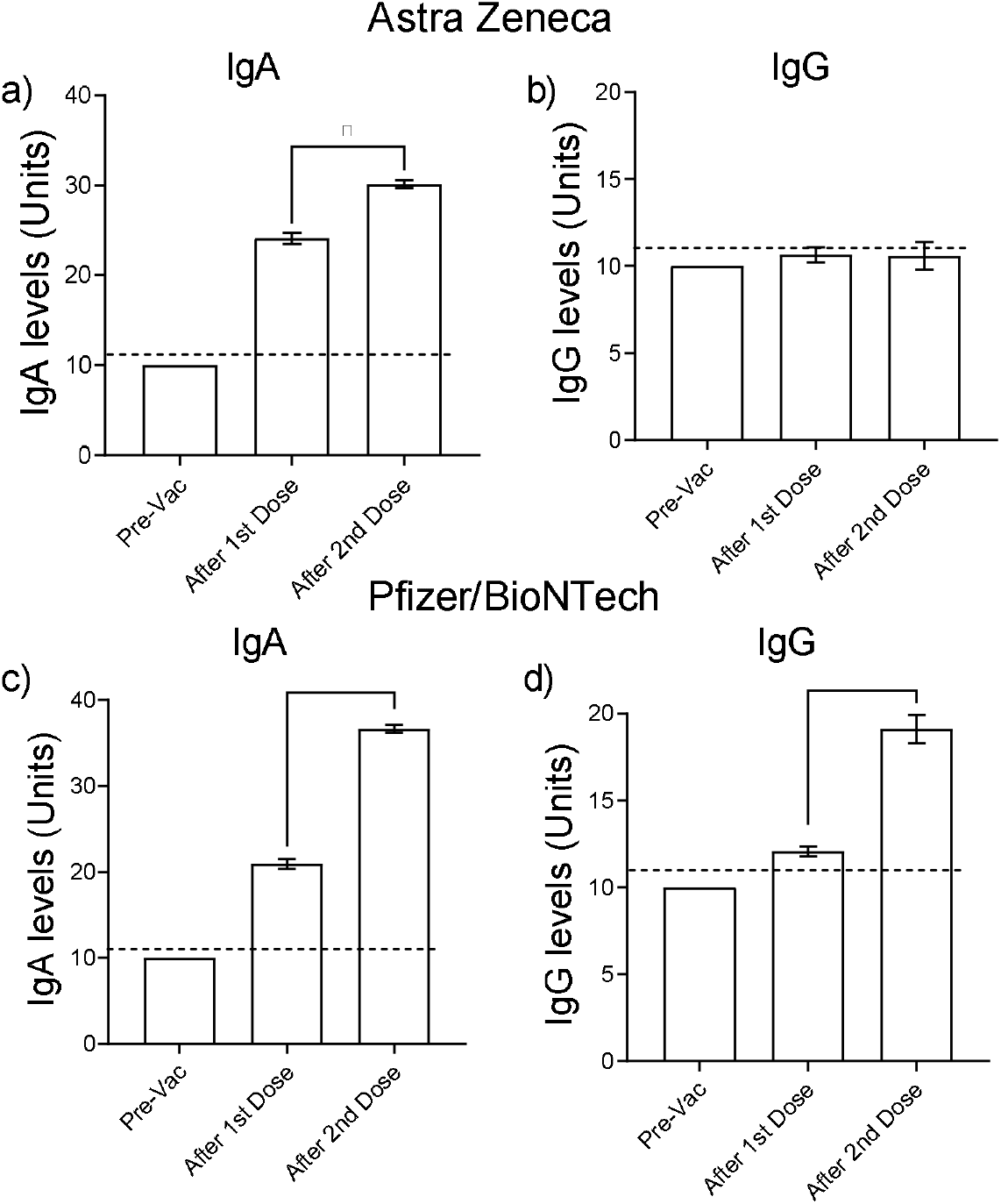
Levels (in Units) of anti-SARS-CoV-2 IgA and IgG in human milk samples before and after vaccination with either AstraZeneca vaccine (a,b) or Pfizer/BioNTech vaccine c,d). Dotted lines: positive cut-off values previously determined for each assay as the mean values of negative control (before vaccination) milk samples. Samples containing >11 U of either IgA or IgG were considered as positive. Asterisks show statistically significant differences between variables (\*\**p* < 0.01; * *p* < 0.05) using the Student’s *t*-test.

All the milk samples taken after the administration of either the Astra Zeneca or the Pfizer/BioNTech vaccine, contained SARS-CoV-2 specific IgA. Moreover, milk samples collected from both women after the administration of the second dose of either of the two vaccines had higher IgA levels compared to those collected after the administration of the first dose (*p*<0.05 for Astra Zeneca and p<0.01 for Pfizer/BioNTech). On the contrary, elevated SARS-CoV-2 specific IgG were detected only in the milk sample collected from the mother vaccinated with the second dose of the Pfizer/Biotech vaccine (**Figure 1**).

### 3.3 Anti- SARS-CoV-2 IgAs and IgGs survive the gastric phase but not the intestinal phase of digestion

The milk sample obtained from the mother vaccinated with the second dose of the Pfizer/BioNTech vaccine was subjected to simulated infant GI digestion and concentrations of anti-SARS-CoV-2 IgA and IgG were determined using the specific ELISA assays (**Figure 2**). Vaccine-induced SARS-CoV-2 IgAs were remarkably stable in the gastric phase but were degraded in the intestinal phase of the simulated digestion protocol (**Figure 2a**). On the contrary, vaccine-elicited anti-SARS-CoV-2 IgGs were degraded in both gastric and intestinal digestion steps (**Figure 2b**).

**Figure 2.**
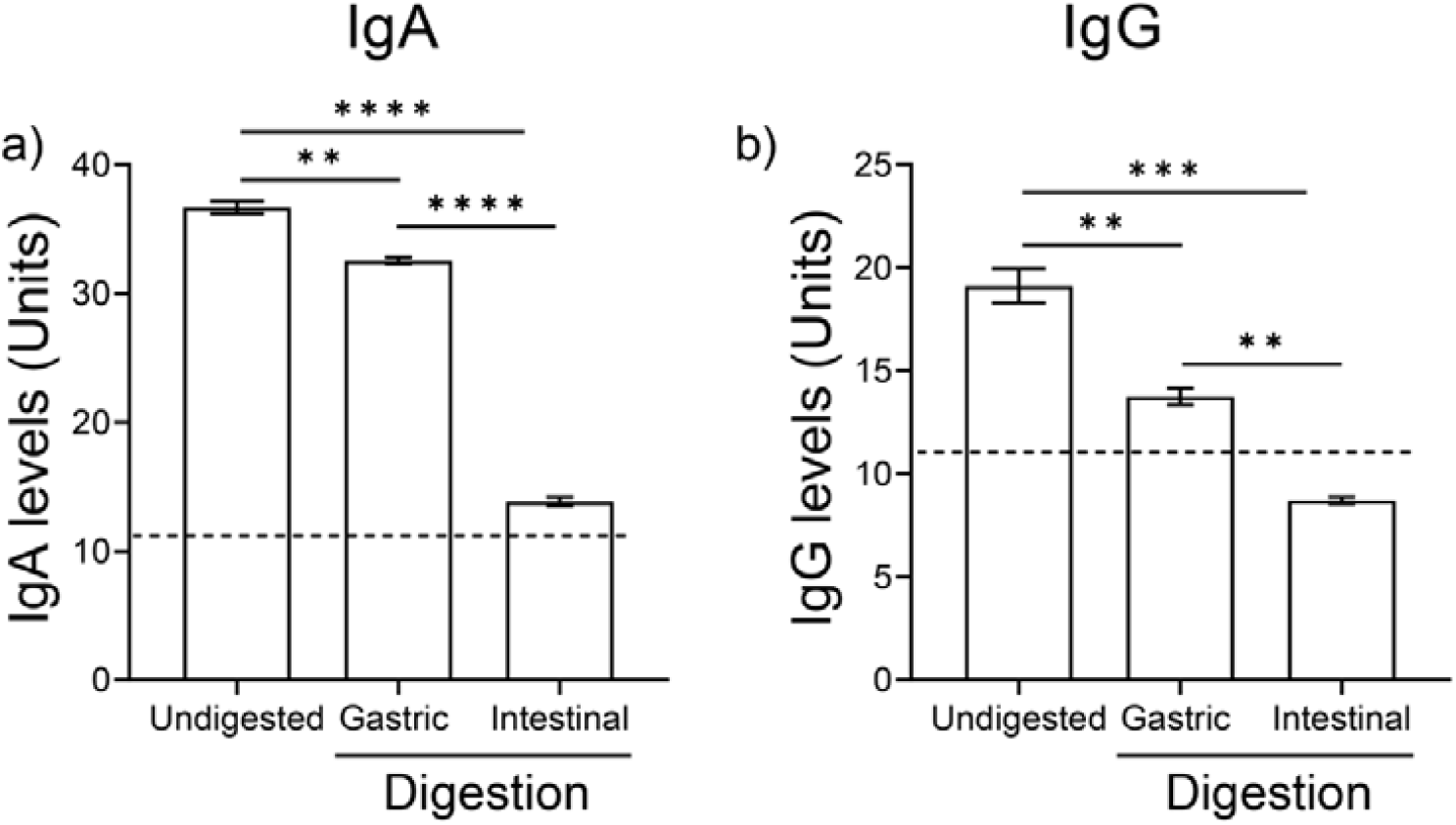
Concentrations of IgA (a) and IgG (b) in milk before and after a 1h-simulated gastric digestion and a 2h-simulated intestinal infant digestion protocol. Dotted lines: positive cut-off values previously determined for each assay as the mean values of negative control (before vaccination) milk samples. Samples containing >11 U of either IgA or IgG were considered as positive. Asterisks show statistically significant differences between variables (** *p* < 0.01, *** *p* < 0.001, \*\*\*\**p* < 0.0001) using the Student’s *t*-test.

## 4. Discussion

In the present study, we demonstrated that commercially available ELISA kits specific for the detection of anti-SARS-CoV-2 IgA and IgG in human serum and plasma are suitable for the detection of the same Igs in human milk samples. Both assays were highly accurate and met the typical validation requirements for their intended use.

Subsequently, using these assays we detected anti-SARS-CoV-2 IgA antibodies in milk following vaccination with both the Pfizer/BioNTech and Astra Zeneca vaccines; however, IgG antibodies were detected only in the milk sample obtained from the mother who received the Pfizer/BioNTech vaccine, with the levels of IgA being higher (>2-fold) compared to that of IgG. IgG antibodies were not detected in the milk from the Astra Zeneca vaccinated individual 3-weeks after the second vaccine dose.

IgA was more abundant than IgG in the two human milk samples tested. This finding does not align with those of a previous study with 10 lactating women who received the Pfizer/BioNTech vaccine and where a IgG-dominant response was observed (Fox et al., 2021). In another study with 6 lactating women who also received both doses of mRNA-based vaccines (Pfizer/BioNTech or Moderna), elevated levels of SARS-CoV-2 specific IgA and IgG antibodies were detected, however an IgG-dominant response was also observed. However, our results are well in agreement with those of a previous study showing an IgA dominant antibody response in the milk obtained from previously infected (with SARS-CoV-2) women (Fox et al., 2020b; Lebrão et al., 2020). Furthermore, in another study of 84 lactating women, milk IgA secretion was observed 2 weeks after vaccination with the first dose of Pfizer-BioNTech vaccine while IgG secretion was observed a week after the administration of the second dose (Perl et al., 2021). In agreement with a previous study (Selma-Royo et al., 2021) our results revealed that while both vaccines increased milk anti-SARS-CoV-2 IgA levels, Pfizer-BioNTech induced higher IgG levels than Astra Zeneca vaccine after administration of the 2^nd^ dose. Moreover, mRNA-based vaccines induced their maximum effect after the administration of the 2^nd^ dose. More studies with higher numbers of participants and various time points are needed to better characterize the immunoglobulin response in milk after both adenovirus and mRNA vaccine technologies for SARS-CoV-2.

Although it has been previously demonstrated that the majority of IgAs in human milk are sIgAs, the ELISA protocol utilised in this study could not determine whether the IgA measured was the secretory type or not. Milk sIgA is thought to be the most important in the infant gut as it neutralizes bacterial and viral pathogens by binding to them, thus reducing their ability to interact with epithelial cells and infect (Demers-Mathieu et al., 2018). However, whether the intramuscular vaccination (IM) elicits the production of sIgA has not been extensively examined. Studies using non-human primates (NHPs) revealed that an IM vaccine may not promote the production of sIgAs (Wilks et al., 2010). Also, this isotype in this conformation is found to be highly resistant to degradation in the GI tract (Fox et al., 2021). Nevertheless, whether the vaccine-elicited IgA are secretory or not is currently under investigation in our laboratory.

Though studies on the production of Igs against SARS-CoV-2 in human milk after infection or vaccination exist (Dong et al., 2020; Fox et al., 2020a; Fox et al., 2021; Yu et al., 2020), we were unable to find previous studies examining the survival of IgA and IgG in the GI system specific for SARS-CoV-2. To neutralize pathogens in the infant gut, maternal milk antibodies must survive digestive protease actions through the GI tract to their site of action. To the best of our knowledge, our case is the first to show that breastmilk SARS-CoV-2 IgA and IgG antibodies survive gastric digestion. Interestingly our results revealed that even though the Pfizer/BioNTech vaccine induced milk IgAs do survive gastric digestion (> 88 %), they are nonetheless fully degraded after the intestinal phase of the *in vitro* digestion protocol utilized in this study.

Few studies have investigated the stability of Igs in digestion. It has been previously demonstrated that gastric digestion my reduce IgGs, but other antibodies including IgAs are not digested in the gastric contents of preterm infants (Demers-Mathieu et al., 2018). Also, a previous study revealed a greater reduction of IgAs compared to IgGs or IgMs in preterm infant stools (Eibl et al., 1988). Two oral supplementation studies (in adults fed bovine colostrum SIgA/IgA, IgM and IgG and in preterm infants fed serum IgA and IgG (Eibl et al., 1988; Roos et al., 1995) demonstrated that IgG and IgM survive intact to the stool, whereas SIgA/IgA does not. However, some studies have demonstrated that human milk-derived SIgA survived intact to the infant stool and urine (Bakker-Zierikzee et al., 2006; Goldblum et al., 1989; Schanler et al., 1986). It has also been demonstrated that overall, the stability of human milk Igs during gastric digestion is higher in preterm infant than in term infants (Demers-Mathieu et al., 2018). In infants, children, and adults, the amount of intact IgG recovered in stool ranges from trace amounts up to 25% of the original amount ingested (Jasion & Burnett, 2015).

A limitation of this study is the fact that it describes a case report of solely two individuals so all results should be interpreted with caution, especially having in mind the great heterogeneity in the immune responses in different individuals (Weaver et al., 1998). We are in the process of obtaining more samples from lactating mothers as the vaccination program in Cyprus progresses. Also, this study did not measure IgA and IgG concentrations in the serum of the lactating mothers, something that could provide interesting information on the comparison of the antibody titers between serum and milk. The use of a single human milk sample to examine the survival of IgGs and IgAs in the simulated GI system also represents a limitation. Furthermore, the current protocol could not discriminate between IgAs and secretory IgAs in the milk samples, something that could be done in the future by employing specific antibodies to the secretory part of the SIgA.

Further studies are needed to elucidate the correlation between IgA/SIgA and IgG levels after vaccination with the age of infant and duration of lactation as well as the nature of vaccine-elicited Igs based on the type of the vaccine. It has been proposed that the immunological profile of mother’s milk is dynamic and can be influenced by several factors including the week of gestation and lactation period (Czosnykowska-Łukacka et al., 2020b). Therefore, the age of the infant and the duration of lactation should also be taken into account when studying Ig profile in human milk post-vaccination.

## Data Availability

The data used to support the findings of this study will be available at the request of the corresponding author.

## Acknowledgements

We are indebted to the milk donors who make this work possible.

